# SARS-CoV-2 infection in vaccinated individuals induces virus-specific nasal resident CD8 and CD4 T cells of broad specificity

**DOI:** 10.1101/2022.05.18.22275292

**Authors:** Joey Ming Er Lim, Anthony T. Tan, Nina Le Bert, Shou Kit Hang, Jenny Guek Hong Low, Antonio Bertoletti

## Abstract

Rapid recognition of SARS-CoV-2-infected cells by T cells resident in the upper airway might provide an important layer of protection against COVID-19. Whether parenchymal SARS-CoV-2 vaccination or infection induce nasal resident T cells specific for distinct SARS-CoV-2 proteins is unknown. We collected T cells from the nasal secretion of COVID-19 vaccinees, who either experienced SARS-CoV-2 infection after vaccination (n=20) or not (n=15) and analyzed their phenotype, SARS-CoV-2 specificity and function. Nasal-resident IFN-γ producing SARS-CoV-2-specific CD8 and CD4 T cells were detected exclusively in vaccinees who experienced SARS-CoV-2 breakthrough infection. Importantly, the vaccine priming of Spike-specific T cells does not suppress the induction of CD8 and CD4 T cells specific for different SARS-CoV-2 proteins (i.e. NP and NSP-12) that persisted in the nasal cavity up to 3 months after infection. These data highlight the importance of viral nasal challenge in the formation of SARS-CoV-2 specific antiviral immunity at the site of primary infection and further define the immunological features of SARS-CoV-2 hybrid immunity.

## Introduction

The human upper respiratory tract is the point of entry and the site of initial SARS-CoV-2 replication(Ahn et al., 2021). Nasal ciliated cells are easily infected due to their high ACE-2 receptor expression and sustain the bulk of initial virus production in vivo(Ahn et al., 2021). In vitro studies have shown that nasal epithelial cells can maintain high levels of viral replication for several weeks despite activation of IFN-a-mediated genes(Gamage et al., 2022) since SARS-CoV-2 can disrupt multiple intracellular antiviral immunity pathways(Banerjee et al., 2020) and block the antiviral efficacy of IFN-a(Xia et al., 2020). Resident T cells that quickly recognize virus-producing infected cells in the nasal cavity can play an important role in rapidly containing and eliminating SARS-CoV-2 infection(Ishii et al., 2022; Zhao et al., 2016), especially after the emergence of Omicron variants of concern (VOC) that elude the preventive efficacy of the neutralizing antibodies induced by current vaccination(Cao et al., 2021).

In animal models of mucosal virus infection, tissue resident virus-specific CD8 T cells act as a first layer of protection. They recognize the virus infected cells and activate innate and adaptive immunity(Schenkel et al., 2014). In infection with respiratory viruses, such as influenza or respiratory syncytial virus, the presence or the adoptive transfer of tissue resident CD8 T cell in the nasal cavity control viral spread and disease severity(Pizzolla et al., 2017a; Kinnear et al., 2018). In Coronavirus infection animal models, protection was dependent on induction of CD4 T cells in the upper airways(Zhao et al., 2016). These airway resident Coronavirus-specific CD4 T cells recruited Coronavirus-specific CD8 T cells by IFN-production(Zhao et al., 2016).

Recent data in mice treated with different vaccine preparations eliciting SARS-CoV-2-specific CD8 or CD4 T cells in the upper airway further confirmed the importance of localized mucosal immunity in SARS-CoV-2 control (Mao et al., 2022; Ishii et al., 2022). However, our knowledge of SARS-CoV-2 T cells in vaccinated or infected individuals has mainly been focused on the analysis of peripheral blood with only few remarkable exceptions.

SARS-CoV-2-specific tissue resident T cells have been observed in human lymph nodes and multiple organs (particularly in the lungs) in SARS-CoV-2 infected convalescent individuals (Poon et al., 2021; Grau-Expósito et al., 2021). T cells specific for SARS-CoV-2 peptides, likely induced by seasonal coronaviruses, have also been detected in the lymphoid tissue of the oral cavity(Niessl et al., 2021) and in bronchoalveolar lavage (BAL)(Maini et al.) of healthy individuals. Finally, a recent analysis of the TCR repertoire of T cells purified from the upper airway of four SARS-CoV-2 infected individuals strongly suggest the presence of Spike-specific CD8 T cells that persisted for up to 2 months after infection(Roukens et al., 2022). Thus, data suggest that SARS-CoV-2 tissue resident T cells exist in the upper and lower airway in infected or healthy individuals, but the impact of parenchymal vaccination or infection in the breath and magnitude of SARS-CoV-2-specific T cells is unknown. Thus, our objective is to evaluate the presence of nasal resident SARS-CoV-2 specific T cells and their functionality and persistence in vaccinated donors who have or have not experienced SARS-CoV-2 breakthrough infection.

## Methods

### Study participants

This study was approved by the SingHealth Centralized Institutional Review Board (CIRB/F 2021/2014). All participant (n=34) were vaccinated with 2 or 3 doses of mRNA vaccine, of which 20 of them have experienced a breakthrough infection identified by COVID-19 rapid lateral flow test. Nasal samples were collected 8 to 149 days (mean=60 days) after last vaccination (Vaccine naïve) and 7-61 days (mean=31 days) after obtaining a negative COVID-19 rapid lateral flow test Convalescent vaccinees). Summarized details on the participants analyzed in this study are listed in Table 1.

**Table 1.** Summarized demographics and vaccination status of donors analyzed in this study.

|  | Naïve vaccinees | Convalescent vaccinees |
| --- | --- | --- |
| **Sample size** | 15 | 20 |
| **Sex** | Female = 6<br>Male = 9 | Female = 9<br>Male = 11 |
| **Age (years)** | 32.2 (24-64) | 32.6 (20-61) |
| **Days post recovery** | NIL | 31.7 (7-61) |
| **Number of nasal lymphocytes retrieved** | $5.4 \times 10^4$<br>($3.2\text{-}9.7 \times 10^4$) | $6.4 \times 10^4$<br>($1.0\text{-}9.7 \times 10^4$) |
| **Vaccination history** |  |  |
| **Number of doses (mRNA vaccine)** | 3-doses = 15 | 2-doses = 12<br>3-doses = 8 |
| **Days post last vaccine** | 60.9 (8-149) | 157.9 (11-382) |

### Nasal sample collection

To collect the nasal lining fluid from individual donors, flocked swab was inserted into the inferior turbinate of the donors and rotated 10 to 20 times. The swab was then placed into a 1mL of AIM-V with 0.1mM of dithiothreitol (DTT; Thermo Scientific) and incubated at 37°C. After 30 minutes, the tube containing the swabs were vigorously vortexed to dislodge the cells. Then, the cells were spun down for washing to remove DTT from the media. The isolated nasal cells were then quantified using flow cytometric analysis and used for subsequent experiments.

### PBMC isolation

Peripheral blood was collected and peripheral blood mononuclear cells (PBMC) from all collected blood samples were isolated by Ficoll-Paque density gradient centrifugation.

Isolated PBMCs were either studied directly or cryopreserved and stored in liquid nitrogen until used in the assays.

### SARS-CoV-2 protein peptide pools

Peptides of 15-mer that overlapped by 10 amino acids spanning the entire protein sequences of NP, Mem, Spike and NSP12 of SARS-CoV-2 were synthesized. Peptides from each protein were pooled into their respective mega-pool and used for subsequent experiments.

### IFN-v El/Spot

Freshly collected nasal cells were stimulated with peptide pools in an IFN-γ ELISPOT assay. Estimated lymphocytes quantity of 5000 to 10000 were seeded per well into ELISpot plates (Millipore Sigma) coated with human IFN-γ antibody overnight at 4°C. Next, the cells were stimulated for 18 hours with the peptide pools at 1 µg/mL. The plates were then incubated with a human biotinylated IFN-γ detection antibody, followed by streptavidin-alkaline phosphatase (streptavidin-AP) and developed using the KPL BCIP/NBT phosphatase substrate (Seracare Life Sciences). To quantify positive peptide-specific responses, spots of the unstimulated wells were subtracted from the peptide-stimulated wells, and the results were expressed as spot-forming cells (SFC) per 10^6^ PBMCs. Results were excluded if negative control wells had more than 30 SFC/10^6^ PBMCs or if positive control wells (anti-CD3/CD28 beads) had less than 100 spots.

### Nasal T cell phenotyping

Cells were re-suspended in phosphate buffered saline (PBS) and stained with Zombie NIR Fixable Viability Kit to exclude dead cells in subsequent analysis. The cells were next washed in FACS buffer with 2mM EDTA, then stained with surface markers anti-CD3-BV605, anti-CD4-BV650, anti-CD8-PE-Cy7, anti-CD69-AF700 and anti-CD103-APC diluted in FACS buffer for 30 minutes on ice. After two more washes in FACS buffer, cells were resuspended in PBS prior to acquisition.

### Phenotyping of SARS-CoV-2 specific T cells

CD3+ cells were depleted from the freshly thawed PBMCs using EasySep Positive Selection Kits II (Stemcell). Flowthroughs consisting of CD3-cells were collected. The cells (5 × 10^5^) were then pulsed with 5µg/mL of peptides or DMSO for one hour in 37°C. After incubation, pulsed cells were washed twice before addition of autologous nasal cells together with anti-CD40L-FITC and anti-CD107a-APC. After 3 hours of incubation at 37°C, the cells were washed in PBS stained with Zombie NIR Fixable Viability Kit to exclude dead cells in subsequent analysis. Then, they were washed in FACS buffer and stained with surface markers anti-CD3-BV605, anti-CD4 PE and anti-CD8 PE-Cy7 diluted in FACS buffer for 30 minutes on ice. After two more washes in FACS buffer, cells were resuspended in PBS prior to acquisition with Beckman Coulter CytoFLEX S analyser.

### Cytokine secretion assay

Freshly isolated nasal cells quantified by flow cytometric analysis and re-suspended in 30uL of culture media (AIM-V + 2% AB serum) or freshly drawn blood was diluted 0.2x with RPMI and stimulated with either 1 µg/mL of peptides or its equivalent concentration of DMSO vehicle. After 16 hours of incubation, the culture supernatant/plasma were collected and stored at -30°C. Cytokine concentrations in the supernatant/plasma were quantified using an ELLA machine with microfluidic multiplex cartridges that measured IFN-γ and IL-2 according to the manufacturer’s protocol. The levels of cytokines present in the supernatant of DMSO controls were subtracted from the corresponding peptide pool-stimulated samples and the values were normalised to 100000 nasal lymphocytes per condition.

## Results

### Characteristics of vaccinated individuals with and without breakthrough infection and collection of nasal cells

We studied the phenotype and SARS-CoV-2 antigen specificities of lymphocytes obtained from the nasal secretion of individuals who were either only vaccinated with BNT162b2 (n=15; indicated as naïve vaccinees) or who experienced a breakthrough infection with Omicron after 2 or 3 doses of BNT162b2 (n=20; indicated as convalescent vaccinees).

Table 1 summarizes the epidemiological characteristics of both groups and the time of nasal sample collection in relation to the last vaccination or infection. Nasal lining fluids were collected using flocked swabs introduced into the inferior turbinate, a method that is well tolerated and does not cause microlesions of nasal mucosa that could result in blood contamination of the specimens(Jochems et al., 2017). The collected swabs were then placed in 1mL of cell culture media with 0.15mM of DTT and vigorously washed to dislodge the cells. The cells were then utilized for phenotypic analysis and different T cell assays (Fig. 1a).

**Figure 1.**
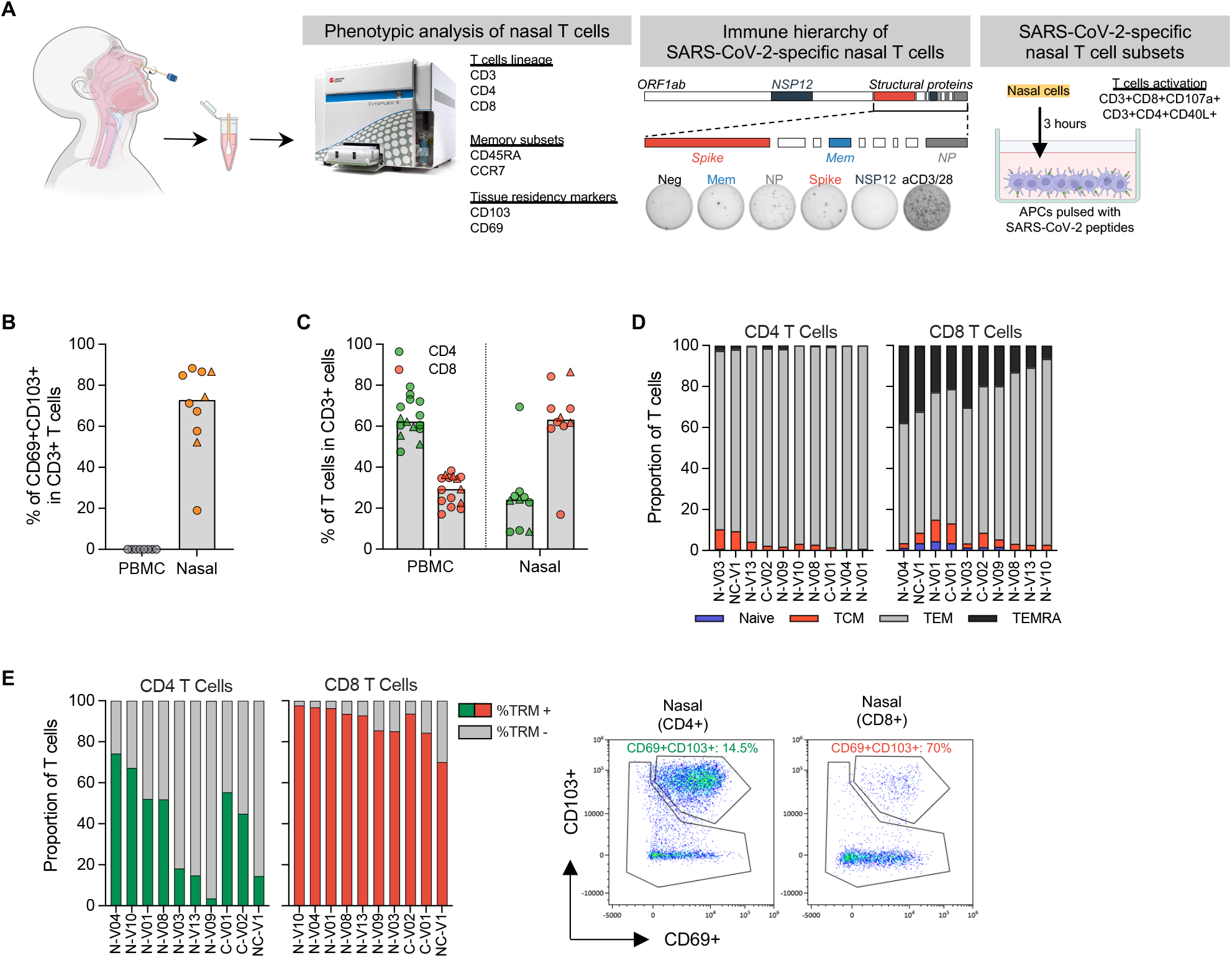
Phenotypic analysis of T cells in nasal secretion. **(a)** Schematic of experimental design; **(b)** Frequency of tissue resident T cells present in PBMC (n=8) and nasal secretion (n=10). Convalescent vaccinees are indicated by a triangle symbol; **(c)** Frequency of CD4 and CD8 T cells present in PBMC (n=14) or nasal cells (n=10). Convalescent vaccinees are indicated by a triangle symbol; **(d)** Proportion of naïve (CCR7+CD45RA+), central (CCR7+CD45RA-; TCM), effector (CCR7-CD45RA-; TEM) and terminally differentiated (CCR7-CD45RA+; TEMRA) memory CD4+ and CD8+ nasal T cells; **(e)** Expression of tissue resident markers in CD8+ and CD4+ nasal T cells (n=10) and corresponding representative plots.

### Phenotype of localized nasal T cells

We first defined the quantity of lymphocytes expressing markers of T cell lineage (CD3), tissue-residency (CD69 and CD103), memory subset (CCR7 and CD45RA) and helper (CD4) or cytotoxic (CD8) identity in 10 individuals of which 3 are convalescent vaccinees (Fig. S1a). This phenotypic analysis demonstrated that CD3+ lymphocytes can be collected from the nasal secretion and that the majority of nasal CD3+ lymphocytes expressed tissue resident markers (Fig. 1b, S1b). Subtyping of the nasal CD3+ lymphocytes showed a predominant CD8+ population unlike that observed in the blood where CD4+ T cells dominate (Fig. 1c, S1c). We also analyzed the expression of CD45RA and CCR7: majority of the T cells were effector (TEM) or terminally differentiated (TEMRA) memory T cells (TEM= CD4+: <90%; CD8+: <60%; TEMRA= CD4+: <5%; CD8+: <20%), while naïve T cells were present at extremely low frequency (Figure 1d, S1d). More importantly, majority (∼90%) of the CD8+ T cells express tissue-residency markers (TRM+), while only ∼40% of the CD4+ T cells are TRM+ (Fig. 1e). There were no observable differences in the phenotype and frequency of nasal lymphocytes between the convalescent and naïve vaccinees.

### Characterization of SARS-CoV-2 specificity of nasal resident T cells

We then analyzed SARS-CoV-2-specificity of T cells collected from the nasal cavity. The cells were activated with peptide pools covering structural proteins able to induce T cell responses (Membrane (Mem), Nucleoprotein (NP) and Spike) in mild/a-symptomatic SARS-CoV-2 convalescent(Bert et al., 2021) and the non-structural protein 12 (NSP12), which induced T cells in individuals with abortive infection(Swadling et al., 2021). We derived the estimated quantity of lymphocytes in each sample by flow cytometry analysis, and we diluted nasal collected cells to perform ELISpot assays with at least 5000 lymphocytes/well. Positive controls obtained by stimulating cells with anti-CD3/CD28 beads and negative controls consisting of unstimulated cells. Data were only analyzed if positive controls showed at least 100 spots. Remarkably, SARS-CoV-2 peptide pools did not elicit any ELISpot response in the 15 naïve vaccinees (13 individuals were negative, 2 individuals with a 1-2 spots), despite presence of Spike-specific T cells in circulation (Fig. 2a-b, S2a). In contrast, SARS-CoV-2 peptide pools activated IFN-γ production by nasal cells in almost all convalescent vaccinees (17 out of 20 showed at least 10 spots/frequency indicated out of 1×10^6^ normalized T cells; Fig. 2a). The requirement of infection for nasal detection of SARS-CoV-2-specific T cells is further supported by data of two vaccinated individuals who were analyzed before and after breakthrough infection where SARS-CoV-2-specific T cells were only detected in the nasal samples after breakthrough infection (Fig. S2b, c).

**Figure 2.**
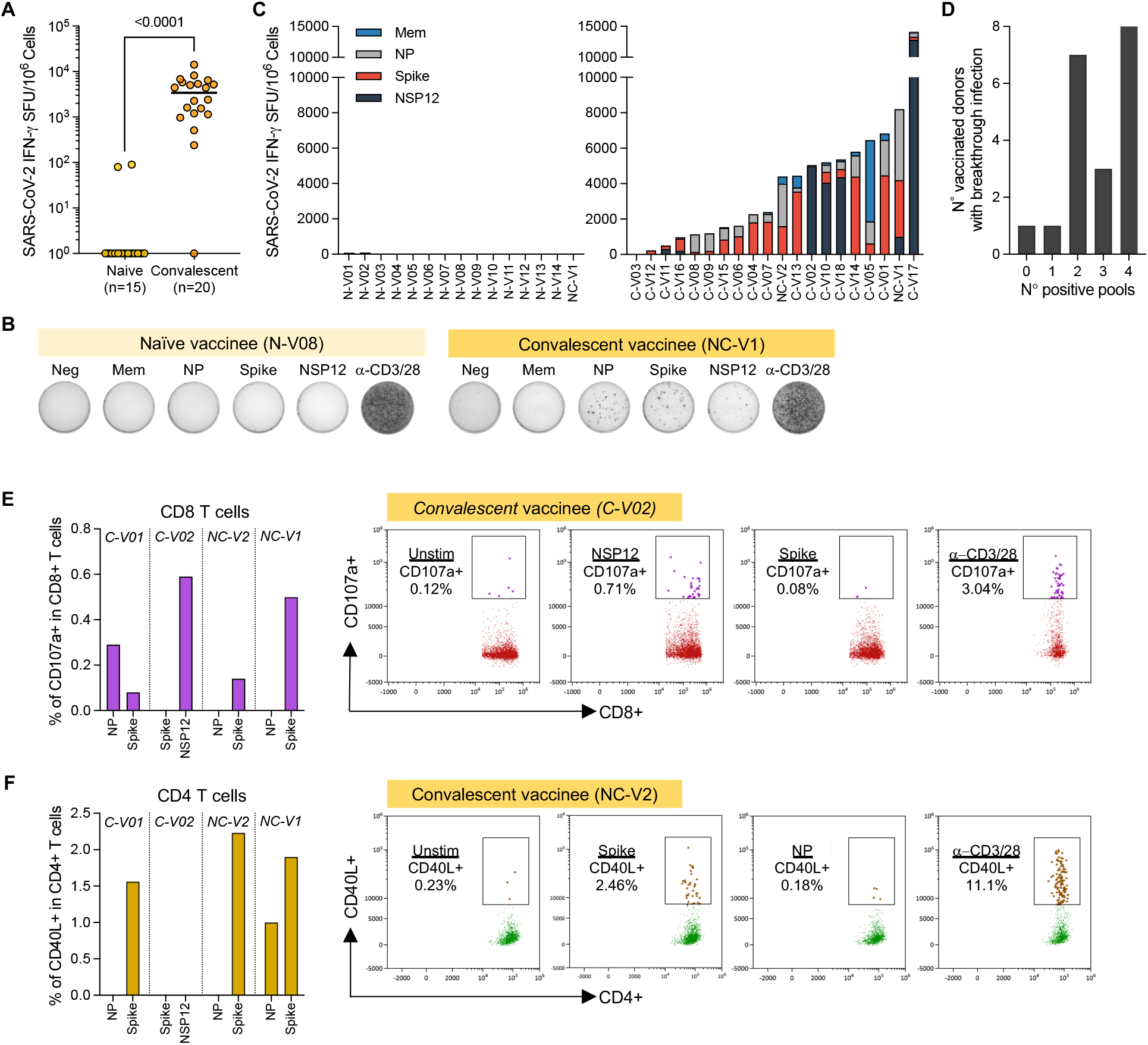
Detection of SARS-CoV-2 nasal resident T cells in convalescent vaccinees but not naïve vaccinees. **(a)** Total frequency of SAR-CoV-2-specific T cell responses in vaccinated without and with breakthrough infection. Lines denote the median value of each group. Each dot represents an individual. Mann-Whitney U test were performed, and the *p-value* is shown in the figure. **(b)** Representative ELISpot wells of nasal cells activated with SAR-CoV-2 peptide pools from a vaccinee without (N-V08) and with breakthrough infection (NC-V1); **(c)** Stacked bars represent the magnitude of SAR-CoV-2-specific T cell responses targeting different peptide pools in vaccinated without (n=15) and with breakthrough infection (n=20). Different colored bars represent indicated SARS-CoV-2 peptide pools.; **(d)** Bar graph shows the number of vaccinated with breakthrough infection (n=20) recognizing the indicated quantity of SARS-CoV-2 peptide pools; **(e-f)** Frequency of (e) CD107a+ CD8 nasal T cells and (f) CD40L+ CD4 nasal T cells detected in four vaccinated donors who experienced breakthrough infection. Corresponding representative plots of frequency of CD107a+CD8+ nasal T cells and CD40L+CD4+ nasal T cells after stimulation with SARS-CoV-2 specific peptide pools.

The response of nasal cells to SARS-CoV-2 peptide pools was heterogeneous in convalescent vaccinees, targeting not only Spike, but also other structural and/or non-structural proteins (Fig. 2c). Spike peptide pool triggered IFN-γ spot formation in 17 out of 20 tested individuals but was dominant only in 8 out of 20. In 4 individuals, nasal lymphocytes recognized preferentially the NP peptide pool, while in 4 others NSP-12 was dominant. In most of the convalescent vaccinees, nasal T cells recognized two to four different proteins (Fig. 2d).

We then characterized in selected convalescent vaccinees whether nasal SARS-CoV-2-specific T cells were CD4+ or CD8+. Nasal samples were incubated for 3 hours with autologous circulating monocytes pulsed or un-pulsed with peptide pools (Fig. 1a). After incubation, we evaluated by flow cytometry the CD3+CD8+ or CD3+CD4+ cells with induced expression of CD107a or CD40L, respectively. Figures 2e and 2f show that nasal T cells responsive to different peptide pools can be either CD8+ or CD4+ T cells. Degranulating CD107a+CD8+ T cells were present in 4 out of 4 convalescent vaccinees who donated additional samples for this analysis. Interestingly, the stronger responses were targeted towards non-Spike antigen, NP and NSP12, from donor C-V01 and C-V02. Nasal CD40L+CD4+ T cells detected from 3 out of 4 individuals, C-V01 and NC-V02, were specific for Spike only, and NC-V1 were targeted for Spike and NP.

### Comparison SARS-CoV-2 T cells in blood and nasal cavity

We analyzed in parallel the ability of nasal cells and PBMCs to recognize the distinct peptide pools (Mem, NP, Spike and NSP12) after breakthrough infection. Figure 3a shows that the profile of antigenic recognition by nasal and circulating T cells was different. As expected, circulating SARS-CoV-2-specific T cells always include a large proportion of Spike-specific T cells. Spike T cell response was dominant in circulating SARS-CoV-2 specific T cells of 8 out of 9 convalescent vaccinees, but such dominance was not always detected in nasal collected T cells. Spike-specific nasal resident T cells were dominant only in 4 out of 9 tested individuals while NSP-12 specific T cells were dominant in nasal T cells in 3 out of 9 and NP-specific T cells in 2 out of 9. Thus, infection causes a hierarchy of SARS-CoV-2 T cell response that is independent from the dominance of vaccine-induced Spike-specific T cells in the blood (Fig. 3a-b).

**Figure 3.**
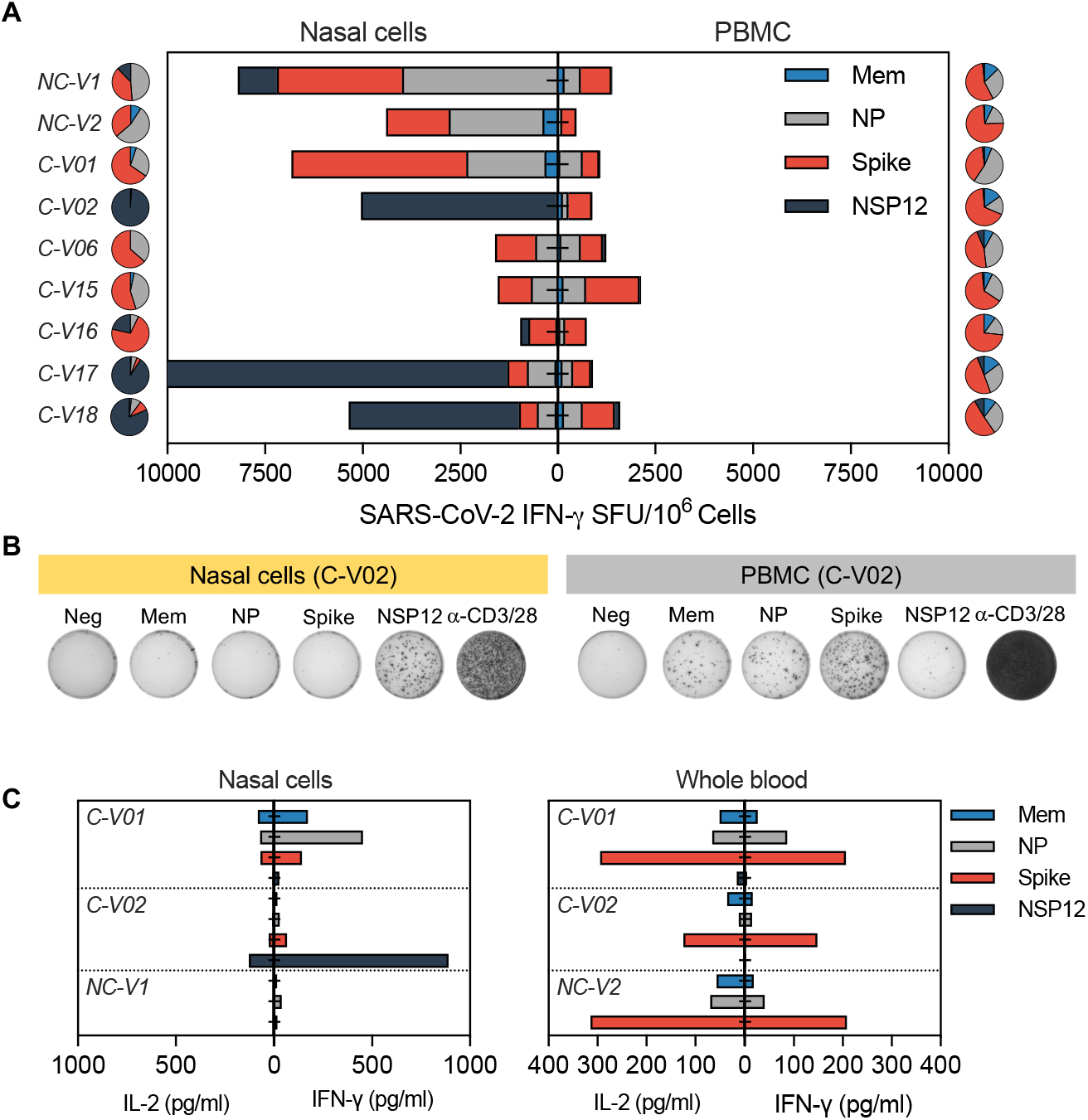
SARS-CoV-2-specific response of nasal and circulating T cells. **(a)** Paired analysis of IFN-γ SFU frequency in nasal cells (left) and PBMC (right) stimulated with SARS-CoV-2 peptide pools of vaccinated (n=9) with breakthrough infection. Pie charts represent the percentage of IFN-γ SFU reactive to the individual SARS-CoV-2 peptides pool; **(b)** Representative ELISpot wells from PBMC and nasal cells; **(c)** Paired analysis of cytokine (IFN-γ and IL-2) secreted in whole blood and nasal cells after stimulation with different SARS-CoV-2 peptide pools.

We also compared the ability of nasal and circulating T cells to produce IFN-γ and IL-2 cytokines after specific peptide stimulation. Supernatants of nasal cells from convalescent vaccinees stimulated with SARS-CoV-2 peptide pools were collected and analyzed for production of IFN-γ and IL-2. An exclusive production of IFN-γ was detected only after peptide pool stimulation, yet IL-2 were barely detectable (Fig. 3c). This profile was different from the cytokine secretion profile of whole blood where high levels of both IFN-γ and IL-2 were present after peptide stimulation.

### Persistence of SARS-CoV-2 T cells in the nasal cavity

We longitudinally evaluated the maintenance of SARS-CoV-2-specific T cells in the nasal environment in 3 convalescent vaccinees. Nasal samples were collected at multiple time points over 3 months after infection and analyzed for the presence of SARS-CoV-2 T cells. We detected SARS-CoV-2-specific T-cell responses at multiple time points for at least 90 days after recovery from infection with no signs of frequency reduction thus far (Fig. 4). These data support the stability of tissue resident T cells in the nasal cavity observed in animal models(Pizzolla et al., 2017a).

**Figure 4.**
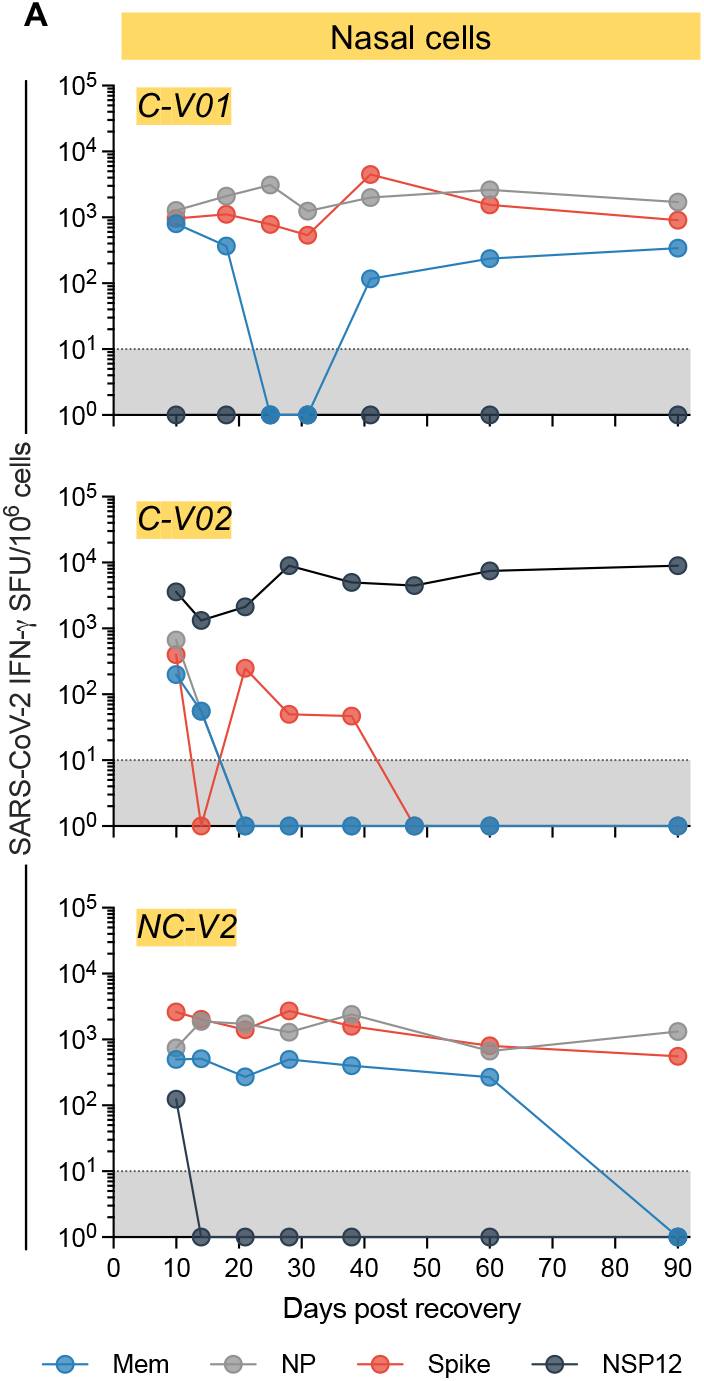
Persistence presence of SARS-CoV-2 specific nasal resident T cells for up to 90 days. Longitudinal analysis of IFN-γ SFU frequency for up to 90 days in nasal cells (n=3). Responses to peptide pools covering different SARS-CoV-2 proteins are indicated by color coding.

## Discussion

We demonstrated that breakthrough SARS-CoV-2 infection of vaccinees lead to the detection of tissue resident CD8 T cells specific for different SARS-CoV-2 proteins in the upper airway. These SARS-CoV-2 multi-specific tissue resident CD8 T cells persist at levels detectable with assays measuring T cell functionality (IFN-γ production by ELISPOT and direct cytokine assay, T cells degranulation) for at least 3 months after infection. On the contrary, identical assays did not detect any T cells specific for Spike or any other SARS-CoV-2 proteins in the upper airway of vaccinated only individuals despite the presence of Spike-specific T cells in their peripheral blood. The ability of infection to recruit a population of T cells specific for different SARS-CoV-2 proteins in the nasal cavity is not unexpected. Spike-specific CD8 T cells after infection have been inferred by analysis of TCR repertoire in the nasal cavity of COVID-19 convalescent patients(Roukens et al., 2022). Furthermore, in animal model of influenza infection virus-specific CD8 T cells are recruited to the upper airway by increased antigen deposition in loco but also by “antigen independent” mechanisms of inflammation mediated by the local infection(Pizzolla et al., 2017a).

Our data do not clarify whether SARS-CoV-2 T cells detected in the nasal cavity of vaccinees after infection were primed in nasal associated lymphoid tissue or in more organized lymphoid structures present in the upper airway (pharyngeal, lingual, palatine tonsils, called the Waldeyer’s ring). Data in animal model suggest that nasal associated lymphoid tissue do not prime virus-specific T cells but support their persistence(Pizzolla et al., 2017b). We were however not expecting the complete absence of SARS-CoV-2 specific T cell response in the nasal secretion of vaccinated only individuals, since cross-reactive SARS-CoV-2 specific CD4 and CD8 T cells were detected in healthy unvaccinated individuals both in pharyngeal tonsils and BAL(Maini et al.; Niessl et al., 2021). For example, there is a clear enrichment of SARS-CoV-2 T cells in BAL as compared to the blood with 0.5-1% of CD4 T cells and 0.1-0.4% of CD8 T cells reported, and these frequencies are numerically compatible with our detection in ELISpot assays. Thus, the absence of SARS-CoV-2 specific T cells in the nasal secretion of vaccinated only individuals show that SARS-CoV-2 infection and not parenchymal mRNA Spike vaccination recruits and retains in the nasal cavity a sizeable population of SARS-CoV-2-tissue memory T cell. These SARS-CoV-2 T cells were likely primed in the organized lymphoid tissue of the upper airway but we cannot certainly exclude that Spike CD8 and CD4 T cells detected in the nasal secretion of our vaccinated convalescent individuals were actually primed in the lymph node draining the site of parenchymal vaccination. This scenario is compatible with the data in mouse model of influenza in whom the nasal cavity has different characteristic of induction, recruitment and persistence of T cells in comparison to the lung(Pizzolla et al., 2017a; b). Another alternative interpretation of our exclusive ability to detect SARS-CoV-2 T cells in the nasal cavity of infected vaccinated individuals might be due to the possible differences in T cell affinity of nasal resident T cells and cross-reactive SARS-CoV-2 T cells detected in the tonsils and BAL of healthy individuals(Maini et al.; Niessl et al., 2021). SARS-CoV-2 cross-reactive T cells can have lower affinity to SARS-CoV-2 epitopes(Bacher et al., 2020) and has been shown to produce lower amount of IFN-γ(Maini et al.; Niessl et al., 2021). We characterized the presence of SARS-CoV-2 T cells by direct stimulation of nasal T cells with peptides pools (without using co-stimulation with anti-CD28 antibodies) and by measuring IL-2 and IFN-γ production. Our method might detect preferentially high affinity SARS-CoV-2 CD8 T cells that produce substantial amount of IFN-γ. Cytokine production of nasal and circulatory SARS-CoV-2 specific T cells showed that nasal resident T cells are secreting high quantity of IFN-γ. A detailed analysis of TCR affinity and transcriptomic profile of SARS-CoV-2 T cells resident in different tissues are needed to define these possibilities.

Irrespective to the causes of the lack of detection of SARS-CoV-2 T cells in the nasal compartment of individuals who were only vaccinated, SARS-CoV-2 breakthrough infection in vaccinees clearly causes a specific accumulation of highly functional SARS-CoV-2 CD8 and CD4 T cells in the nasal cavity. These cells might quickly trigger a localized secretion of the antiviral IFN-γ cytokine after antigen encounter and start a chain of innate and adaptive immune events able to control viral replication, as shown in different mice models of viral infection(Zhao et al., 2016; Pizzolla et al., 2017a; Schenkel et al., 2014; Mao et al., 2022; Kinnear et al., 2018; Ishii et al., 2022).

Despite the limited sample size of this study, our analysis of cellular immunity in nasal secretion complements the growing experimental evidence showing that only infection after vaccination but not parental vaccination alone induces a mucosal SARS-CoV-2-specific antibody response characterized by robust production of secretory Spike-specific IgA(Isho et al., 2020; Sano et al., 2021; Azzi et al. 2022; Terreri et al., 2022; Collier et al., 2022). Our data also provide further evidence of the peculiarity of anti-SARS-CoV-2 immunity in individuals with “hybrid” immunity. The combination of SARS-Cov-2 infection and vaccination elicit stronger antibody and T cell response in the blood(Reynolds et al., 2021) and a broad repertoire of anti-Spike specific B cells(Terreri et al., 2022; Rodda et al., 2022) and a functional profile of circulating CD4 T cells characterized by production of IFN-γ and IL-10(Rodda et al., 2022). This T cell cytokine profile is associated with virus-respiratory protection in animal models(Zhao et al., 2016) and with a/mild-symptomatic SARS-CoV-2 infection in humans(Bert et al., 2021; Grau-Expósito et al., 2021). Breakthrough infection has also been shown to expand the CD8 T cell repertoire in the circulation(Minervina et al., 2022) and our characterization of SARS-CoV-2 specific T cells in the nasal cavity demonstrated that an expansion of CD8 and CD4 T cells specific for different antigens is not an exclusive feature of circulating peripheral blood. Taken together these immunological features can explain why individuals with “hybrid” immunity can control SARS-CoV-2 Omicron replication quicker that vaccinated only(Marking et al., 2022) and why a superior immune protection is induced by infection over mRNA vaccination alone(Chemaitelly et al., 2022).

The observation that SARS-CoV-2 infection caused an enrichment in the nasal cavity of tissue resident CD8 T cells specific not only for Spike but also for different structural and/or non-structural viral proteins can also provide better recognition of Omicron variants(Marking et al., 2022). Since the NSP-12- and NP-specific CD8 T cells are less likely to be affected by the AA-mutations preferentially present in the Spike protein of Omicron BA.1 and BA.2.

Finally, the persistence of tissue resident SARS-CoV-2 T cells with no experimental sign of frequency reduction in the first 90 days after SARS-CoV-2 clearance is in line with mice data showing that virus-specific nasal T cells display minimal decay overtime(Pizzolla et al., 2017a). Therefore, nasal resident SARS-CoV-2 T cells might represent a long protective immune cell population. Further longitudinal analysis will be necessary to test whether such SARS-CoV-2 nasal resident T cells can persist for years as we have seen in circulating memory T cells after SARS-CoV-1 infection(Le Bert et al., 2020).

There are limitations in this study; in addition to the small sample size, phenotypic analysis of the nasal T cells and analysis of their SARS-CoV-2 specificity was limited to few markers of tissue residency (CD69 and CD103) and memory (CD45RA and CCR7) and to selected SARS-CoV-2 proteins (NP, membrane, Spike and NSP-12). The limited number of T cells collected impose these limitations. To better define the breath of the nasal resident T cell response against SARS-CoV-2 it will be necessary to analyze their ability to recognize epitopes derived from others structural, non-structural proteins or out-of-frame Open Reading Frames that have been shown to induce robust CD8 T cell response in circulation(Weingarten-Gabbay et al., 2021). The potential distinct functionality of Nasal SARS-CoV-2 T cells will need also to be analyzed by analysis of their transcriptomic profile, in addition their ability to produce IFN-γ and degranulate. Finally, as already mentioned, study of the durability of the nasal resident SARS-CoV-2 T cells over 3 months are warranted to understand the long-term impact of nasal resident T cells in SARS-CoV-2 protection.

## Data Availability

All data produced in the present work are contained in the manuscript

## Figure legends

**Supplementary Figure 1.**
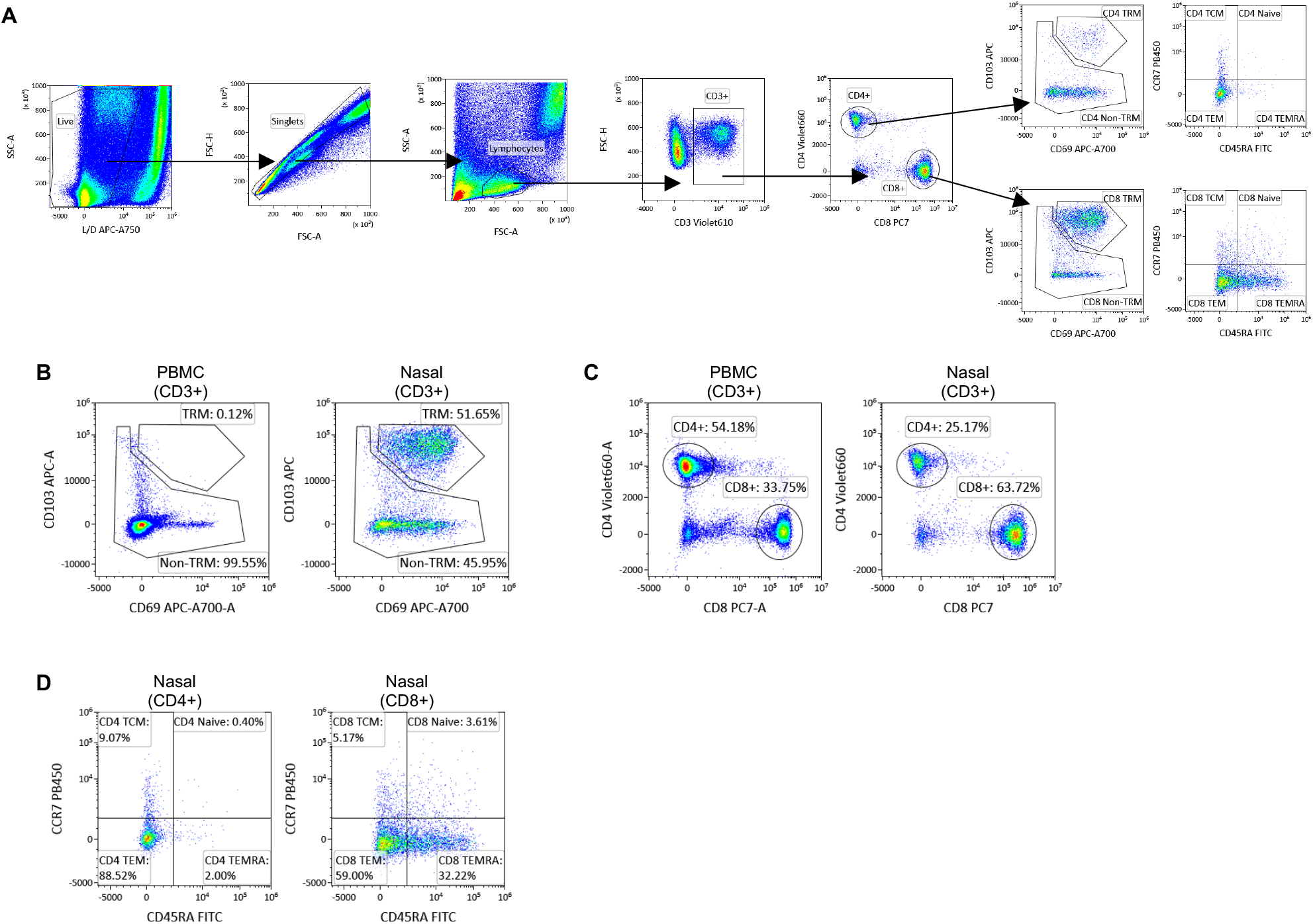
Flow cytometric analysis of nasal T cells. **(a)** Representative gating strategy of nasal T cells phenotypic analysis; **(b)** Representative flow plots of expression of tissue residency markers in PBMC and nasal cells; **(c)** Representative flow plots of expression of proportion of CD4 and CD8 T cells present in PBMC and nasal cells; **(d)** Representative flow plots of expression of proportion of memory subsets in CD4 and CD8 T cells.

**Supplementary Figure 2.**
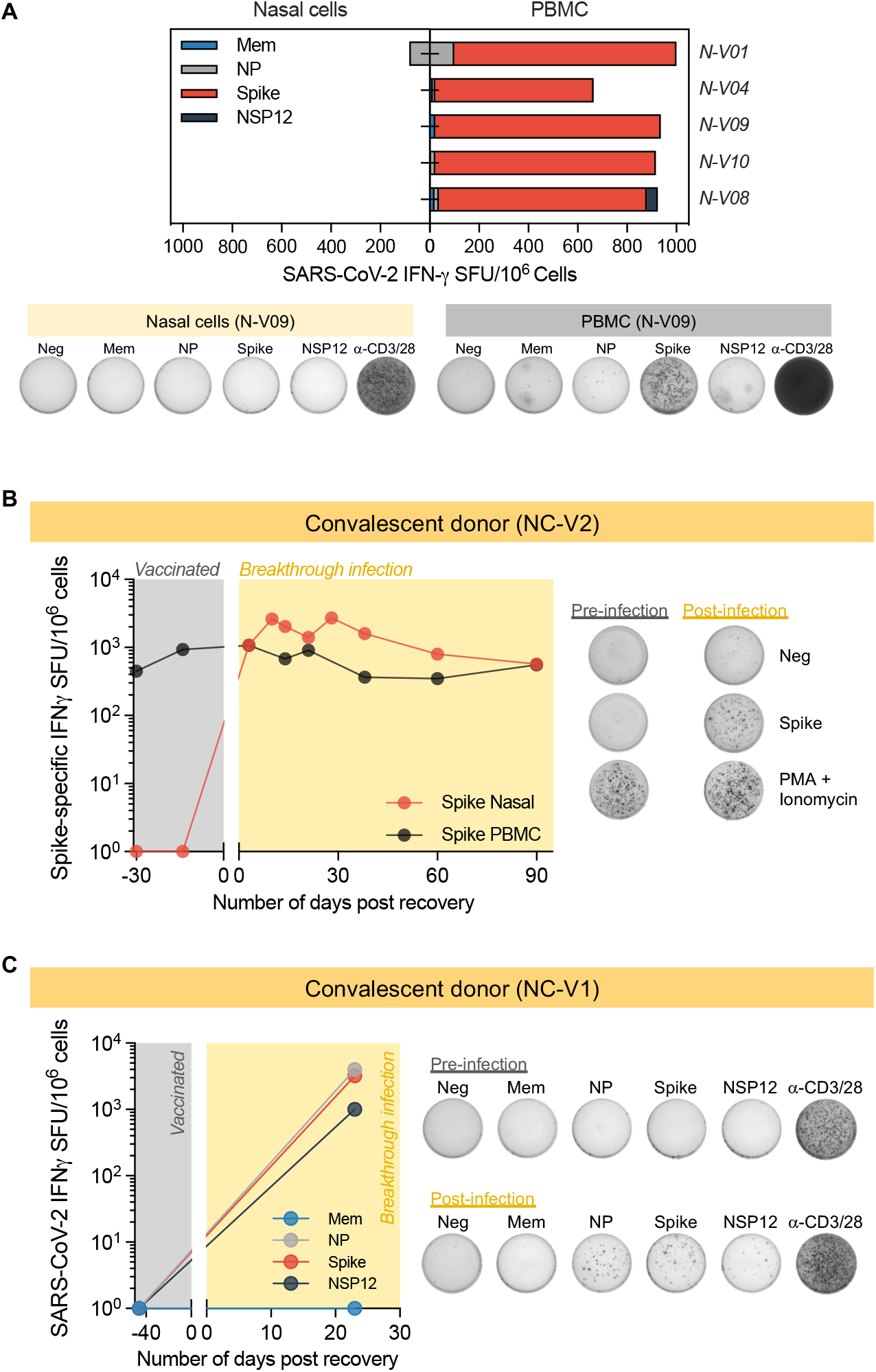
Representative ELISpot wells showing SARS-CoV-2 specific T cells in naïve and convalescent vaccinees. **(a)** Paired analysis of IFN-γ SFU frequency in PBMC and nasal cells stimulated with SARS-CoV-2 peptide pools of vaccinated (n=5) with breakthrough infection. Pie charts represent the percentage of IFN-γ SFU reactive to the individual SARS-CoV-2 peptides pool. Representative ELISpot wells from PBMC and nasal cells; **(b-c)** Longitudinal analysis of Spike-specific IFN-γ SFU frequency present in PBMC and nasal cells of a vaccinated individual before and after breakthrough infection. Representative ELISpot wells of (b) Spike-specific and (c) SARS-CoV-2-specific T cell response from vaccinated donor before and after breakthrough infection. Representative ELISpot wells of T cell response from vaccinated donor before and after breakthrough infection.

**Supplementary Figure 3.**
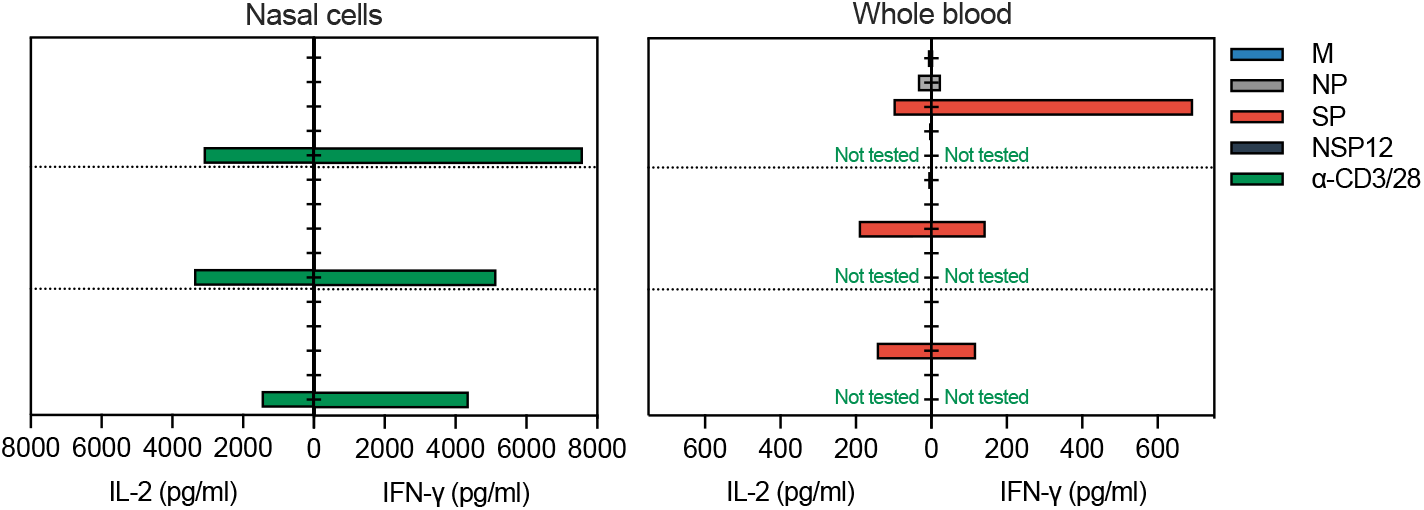
Paired analysis of cytokine (IFN-g and IL-2) secreted in whole blood and nasal cells after stimulation with different SARS-CoV-2 peptide pools of vaccinated donors (n=3).

## Notes

### Competing Interest Statement

N. Le Bert and A. Bertoletti reported a patent for a method to monitor SARS-CoV-2-specific T cells in biological samples pending.

### Funding Statement

This study was funded by the Singapore Ministry of Health National Medical Research Council under its COVID-19 Research Fund COVID19RF3-0060, COVID19RF-001 and COVID19RF-008.

### Author Declarations

The study protocol was reviewed and approved by SingHealth Centralized Institutional Review Board (CIRB/F 2021/2014). All donors provided written consent for enrolment

